# Evaluating the impact of compound heterozygosity involving microdeletions and sequence-level variants: findings in autism

**DOI:** 10.1101/2025.10.17.25338215

**Authors:** Worrawat Engchuan, Kara Han, Rayssa MMW Feitosa, Nelson Bautista Salazar, David J Mager, Shania Wu, Faraz Ali, Alexander Chan, Marla Mendes de Aquino, Xiaopu Zhou, Rulan Shaath, Nickie Safarian, Bhooma Thiruvahindrapuram, Thomas Nalpathamkalam, Giovanna Pellecchia, Jill de Rijke, Mehdi Zarrei, Elemi Breetvelt, Stephen W. Scherer, Brett Trost, Jacob Vorstman

## Abstract

Compound heterozygous events involving a chromosome deletion and on the remaining allele a functional DNA sequence-level variant can underpin a range of medical conditions. Most large-scale genetic studies do not include a systematic analysis of such compound heterozygous deletion (DelCH) events. We developed three frameworks: i) traditional burden analysis; ii) deletion-matched burden analysis; and iii) transmission disequilibrium test (TDT), to examine the possible contribution of DelCH to clinical presentations, and report results of their implementation in 9,766 families of autistic individuals. Across the three strategies, we observed enrichment of rare DelCH events in autistic individuals at a nominal significance level for individual tests. Collectively, six genes; *CFHR4*, *HSDL1*, *MYO15A*, *NEFH*, and three olfactory receptor genes; *OR1A2*, *OR4P2*, were affected by DelCH events in at least two unrelated autistic individuals (and not in unaffected family members), while the reverse analyses identified no genes (p<2.2 x 10^-16^). Gene set enrichment analysis of the extended network of candidate genes showing a remarkable convergence to processes related to neurogenesis. Our findings suggest a modest role for DelCH events in ASD. The strategies described here are available via a GitHub repository, allowing the research community to examine the role of DelCH in other genome sequencing cohorts.

## 1. Introduction

Many biological processes require finely tuned regulation of gene expression, in which the expression level of each gene (or gene pair) must fall within a certain range^1,2^. Genomic copy number variants (CNVs), which include deletions and duplications of DNA, can impact biological function by decreasing or increasing gene dosage. CNVs can be pathogenic if they affect expression of genes that are sensitive to dosage changes^3^.

Genes for which just one copy is insufficient to maintain normal biological function are described as ‘haploinsufficient’. The estimated number of human protein-coding genes that are haploinsufficient is just under 3,000^4^, based on a gnomAD (v2.1)^5^ loss-of-function observed/expected upper bound fraction (LOEUF) threshold of below 0.35^6^. Consequently, for most human genes, one copy is sufficient for normal biological function^7–10^. However, pathogenicity may still arise when the remaining copy of a deletion-impacted gene is also affected by functionally relevant variation^11^. The scenario in which both alleles of a given gene are impacted by different deleterious genetic variants is called compound heterozygosity. In the present work, we focus on an understudied type of compound heterozygosity, namely the co-occurrence of a deletion on one allele and a sequence-level variant (single nucleotide variant [SNV] or indel) affecting the other allele, hereafter referred to as deletion compound heterozygosity (DelCH).

Examining DelCH events in disease cohorts is important for several reasons. First, DelCH events are typically under-scored in large-scale genomics studies and thus including them should contribute to achieving a more comprehensive library of disease-associated genetic variation. Second, DelCH may help explain why some individuals with a clinical phenotype have potentially clinically relevant deletions inherited from an unaffected parent. Third, including DelCH in genetic analyses may lead to the identification of recessive disease genes that would otherwise escape detection. Although less frequent than compound heterozygosity caused by sequence-level variants on both alleles, case reports of diseases caused by DelCH have been reported. These include phenotypes ranging from osteogenesis imperfecta to chylomicronemia, often involving inheritance of the deletion from one parent and the sequence-level variant from the other, with both parents phenotypically unaffected^12–15^. There are also several studies reporting neurodevelopmental and/or psychiatric phenotypes putatively caused by this genetic mechanism^16–20^. Beyond these case reports, systematic analyses of this phenomenon in case-control cohorts are scarce, and include two studies providing tentative evidence for a role for DelCH in schizophrenia^21^ and autism spectrum disorder (ASD)^22^.

ASD is a neurodevelopmental disorder whose common features include social/communication impairments, repetitive behaviors, and restricted interests^23^. The severity of these characteristics varies greatly—some autistic individuals require high levels of support in their daily lives, whereas others are entirely independent and excel in many societal domains. In addition to its phenotypic variability, studies indicate a high level of genetic heterogeneity. Over the past three decades, the proportion of autistic individuals for whom a contributing rare genetic variant can be identified has increased from approximately 2-4% to approximately 5-25%, depending on the genotyping method and the characteristics of the population examined^24^. The identification of genetic variants associated with ASD is a crucial step toward elucidating the genes and biological mechanisms underpinning this condition^25^. For these reasons, genomics research in ASD has focused on the analysis of large cohorts and the evaluation of different genetic variant types. As a result of these efforts, the number of ASD-associated genes continues to rise, with over one hundred genes now definitively implicated in ASD etiology^26–28^. It is also noteworthy that a subset of these genes was identified through the study of recessive mutations being found in mostly consanguineous families^29,30^.

Variants that substantially affect ASD susceptibility are always rare and include sequence-level variants in ASD-associated genes as well as structural variants such as CNVs, which often affect more than one gene. Common variants individually exert small effects, but in aggregate have larger effects and account for a substantial fraction of ASD liability^31^, although ASD polygenic scores currently explain only about 2.5% of its variance^32^. Increasing efforts are being made to understand the combined influence of common and rare variants^33–35^, through the inclusion of the most comprehensive catalogue of variants detectable in the human genome^28^. There is an increasing realisation that an integrative approach to identifying genotype-phenotype associations, considering *all classes* of genetic variation, is a crucial step towards precision medicine, both in psychiatry and across all medical disciplines^36^. The reliable identification of compound heterozygous events is part of such an integrative approach to genetic analysis.

Unfortunately, analytical strategies for examining DelCH on a cohort-wide scale have not yet been developed. There are several reasons for this. First, this analysis requires whole-genome sequencing (WGS) data, which only became available for researchers in recent years. Furthermore, given that the mechanism typically consists of two *a priori* rare events at the same locus, the role of DelCH in any given condition is likely modest, suggesting that a large sample size is necessary to achieve sufficient statistical power. In addition to this “lightning striking twice” feature, the data preparation is demanding, as each individual presents with unique deletion regions which must be examined individually for sequence-level variants on the other allele. In addition, the selection of variants requires careful consideration; for example, deletions or deleterious sequence-level variants that are known to affect ASD susceptibility in an autosomal dominant fashion are not candidates for DelCH, since they do not require the second hit on the other allele; consequently, patients with dominant pathogenic variants associated with ASD are *a priori* excluded from the analysis. Finally, defining exactly where on the non-deleted allele the second variant can occur can be challenging, as it depends on the type of data and chosen analytical approach; it can be based on the start and end positions of the deletion, or on the boundaries of any gene(s) affected by the deletion (Fig. 1). The selection criteria for these variables require careful consideration as they impact the results of the analysis.

**Fig. 1.**
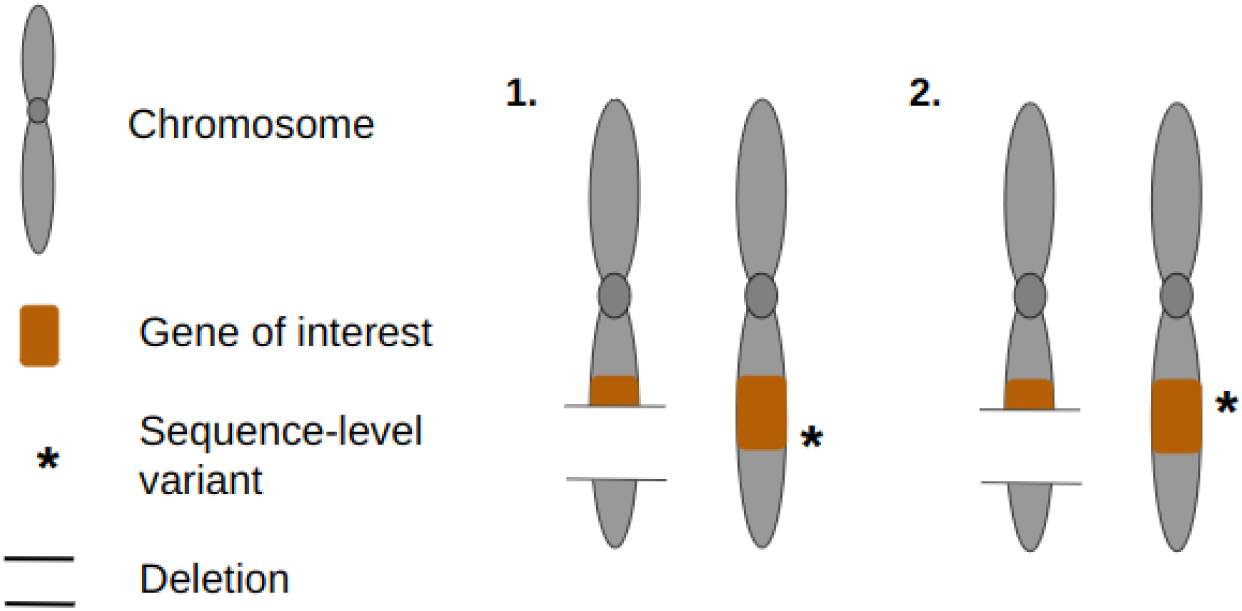
Examples of compound heterozygous events involving a deletion on one allele and a sequence-level variant on the other allele. Scenario 1: The coding region of a gene (orange) is partially deleted on one allele, with a sequence-level variant (asterisk) on the other allele that is within both the coding region and the boundaries of the deletion. Scenario 2: same as scenario 1, except the sequence-level variant is outside the boundaries of the deletion.

In this study, we developed three complementary strategies for examining the role of genome-wide DelCH in large cohorts of individuals with WGS data, namely, (1) traditional burden analysis, (2) deletion-matched burden analysis and (3) transmission disequilibrium test (TDT) (Fig. 2). While our approaches can be applied to any condition with a putative genetic contribution, we evaluated our methods in a large WGS dataset of 9,766 families of autistic individuals from the MSSNG^28,37^, Simons Simplex Collection (SSC)^28,37,38^, and SPARK^39^ cohorts. Along with a discussion of the advantages and disadvantages of each approach, we make the analytical pipeline available to the scientific community via a GitHub repository for further improvements and application in other cohorts. Ultimately, we also analysed and discussed the significance of our results in the tested cohorts.

**Fig. 2.**
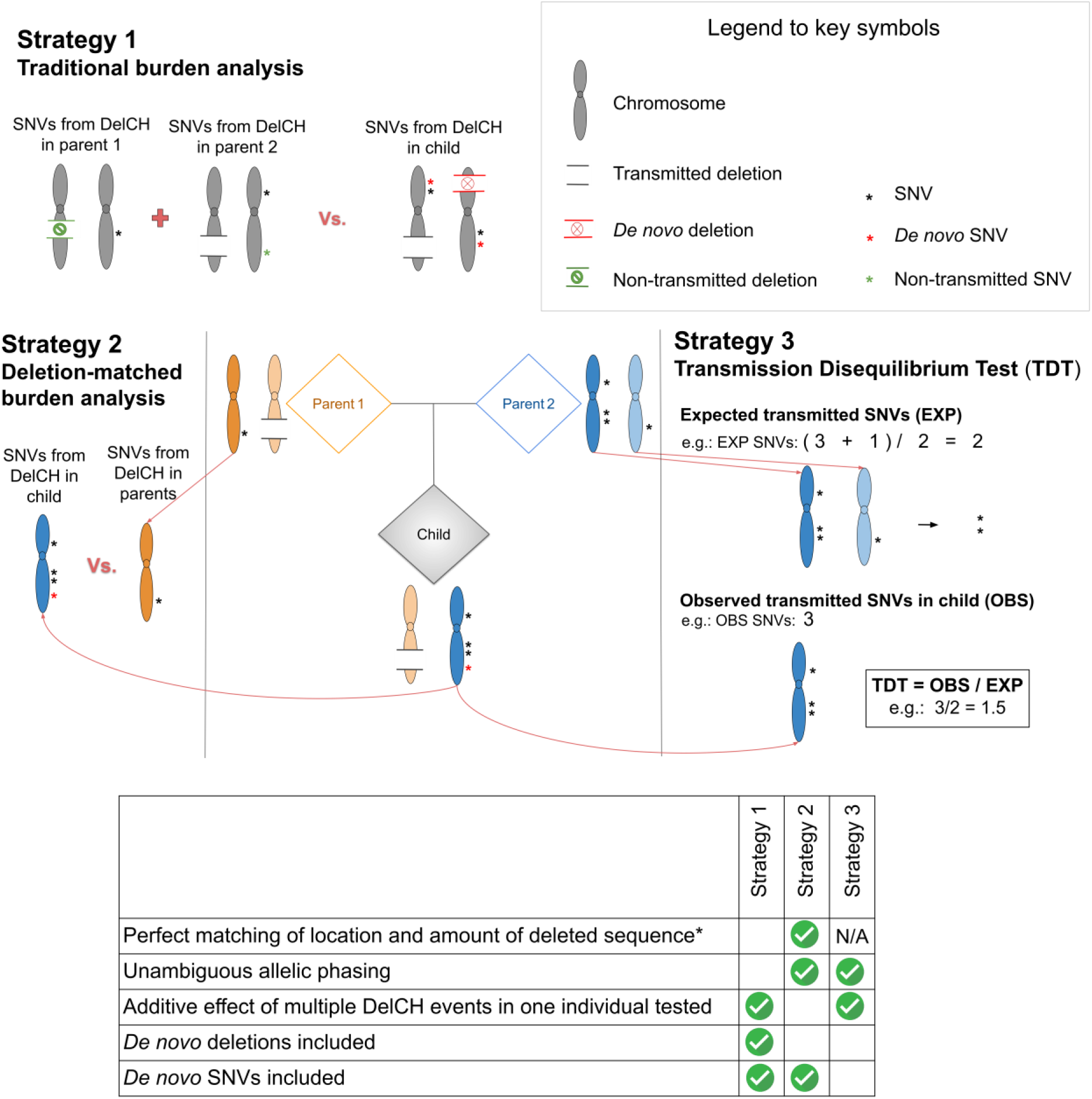
The three strategies used to examine the role of DelCH events in disease. **Strategy 1** (“traditional burden analysis”) is a burden analysis of DelCH events, comparing children and parents, but agnostic to inheritance status of the variants involved (i.e., inherited, non-transmitted, and *de novo*). **Strategy 2** (“deletion-matched burden analysis”) compares the burden of DelCH events in inherited deletions between children and their deletion-transmitting parents. **Strategy 3** (“TDT”) is a transmission disequilibrium test, which is calculated using the expected and observed inherited SNVs in the non-deletion-transmitting parent. Note that the Expected SNVs (EXP) is the total SNVs of non-deletion-transmitting parents divided by two (diploid genome). For TDT, children can either be disease-affected children alone, or (as implemented here) a comparison between transmission to affected children and transmission to unaffected siblings. DelCH = deletion compound heterozygosity; SNV = single nucleotide variant; Vs = versus. * For traditional burden analysis, the comparison is between children and both parents. For deletion-matched burden analysis, children are compared to the parent from whom the deletion was inherited. This does not apply to TDT, as the comparison was not done between deletion-carriers.

## 2. Methods

### 2.1. Data filtering

Short-read WGS data were obtained from three large cohorts of ASD families: MSSNG^28,37^, SSC^38^, and SPARK^39^. A total of 9,766 families were analyzed in this study. SNVs were detected using the HaplotypeCaller from Genome Analysis Toolkit (GATK^40^), while deletions were detected using two structural variation detection pipelines: a read-depth based pipeline (CNVnator^41^ and ERDS^42^) and a paired-end mapping-based pipeline (Manta^43^ and DELLY^44^). Standard quality controls were performed as described previously^28^. We only included SNVs in coding sequence (annotated by ANNOVAR^45^ as “exonic” or “exonic; splicing” in the typeseq priority column) and those affecting canonical splice sites (i.e., an SNV affecting the 2 bp at either end of an intron). Given that DelCH follows a recessive mode of action, the X chromosome was excluded from our analysis. Given that paired-end mapping-based pipeline may also detect somatic deletions, we removed deletions encompassing at least one high-quality heterozygous SNV call, indicative of likely mosaicism. The remaining germline deletions were those detected by either of the two pipelines, with breakpoints defined by the read-depth-based pipeline if a deletion was detected by both.

Gene definitions were obtained from RefSeq release 200 (GRCh38), which contains 19,433 protein-coding genes^46^. The recessive mode of action of DelCH events implies that each component (the deletion and the sequence-level variant) is not expected to be pathogenic on its own. Therefore, we excluded 1,149 families where the ASD phenotype was already attributable to recurrent CNVs, large deletions (>3Mb), or dominant loss-of-function SNVs or indels (i.e., stop gain, frameshift indels, canonical splicing variants) impacting 134 previously reported ASD candidate genes^28^. Only trios/quad families (n=6,902 families) were retained for subsequent analyses to ensure ability to accurately infer deletion and SNV phasing. Furthermore, we excluded genes predicted to be of dominant effect/loss-of-function intolerant (i.e., those with gnomAD v2.1 LOEUF score < 0.35), retaining 16,636 genes. Of these, 8,036 genes were impacted by at least one deletion with a frequency of less than 10% in our dataset. This gene set was the substrate for our main analysis (Fig. 3).

**Fig. 3.**
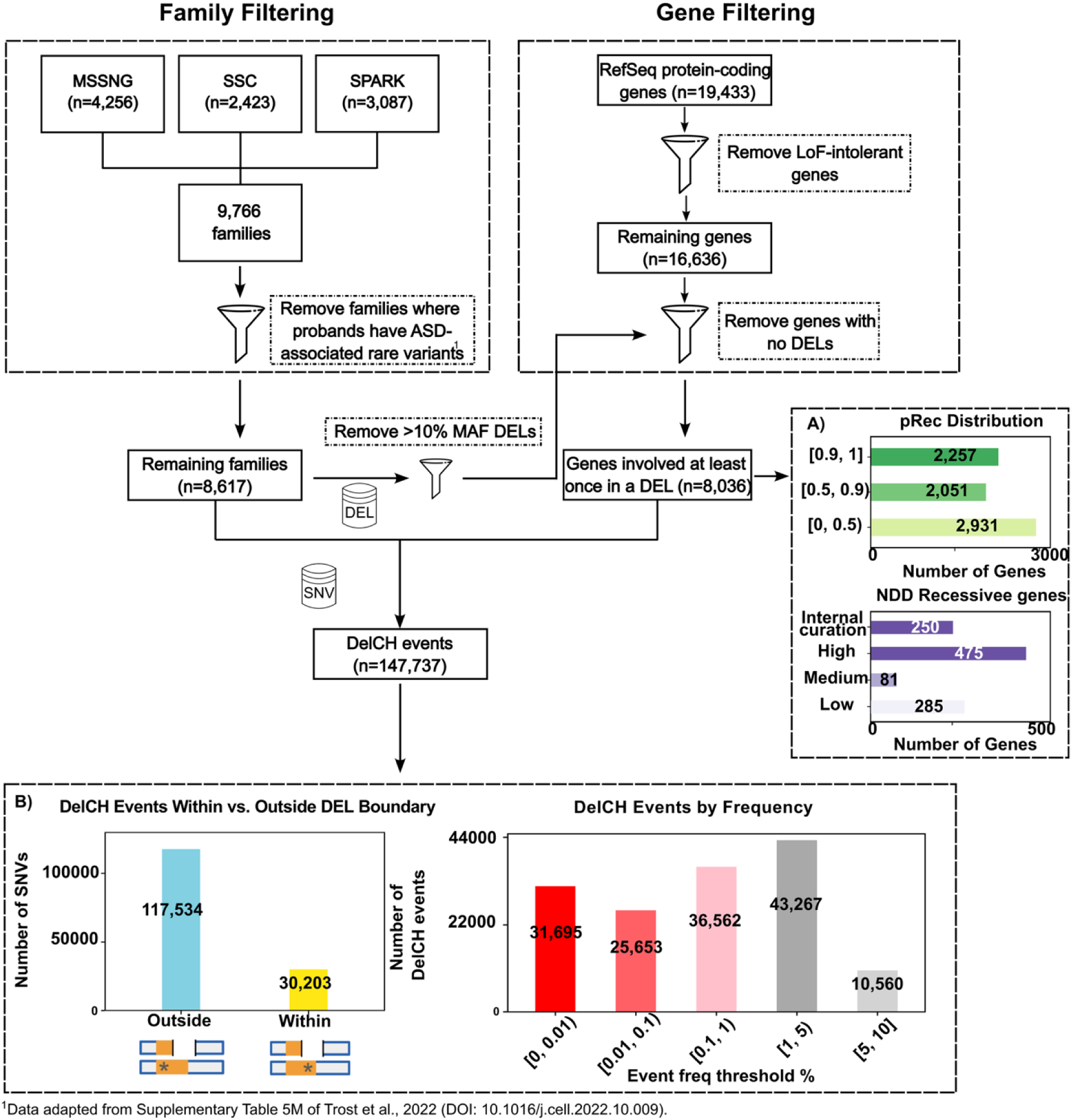
Filtering procedure for family, gene, and variants. Families: we retained quad or trio families where no ASD-relevant variants were identified in any of the family members. Genes: we retained only protein-coding genes not defined as LoF-intolerant by gnomAD LOEUF cut-off of <0.35). Variants: Exonic (and splicing) SNVs across all MAF range and deletion with MAF < 10% were retained for our analyses. **A)** pRec and neurodevelopmental disorder (NDD) recessive gene distributions for genes involved in at least one deletion. Low, Medium, and High are based on Genomic England’s neurodevelopmental disorder recessive genes set, while *Internal curation* refers to our curated neurodevelopmental genes set **(supplementary section 1, table S1)**. **B)** Distribution of DelCH events within versus outside deletion boundaries and by frequency. The asterisk represents an SNV on the non-deletion-harbouring allele inside the coding region of a gene (orange bar).

#### 2.1.1. Annotation for recessive intolerance and association with neurodevelopmental conditions

We then annotated these 8,036 genes by the gnomAD v2.1^5^ pRec score, which predicts the probability of intolerance to biallelic loss-of-function variants, but not to heterozygous loss-of-function variants. Of these, 7,239 genes had pRec scores available, and 2,257 had high pRec scores, the latter indicating a low tolerance to biallelic loss-of-function variants (Fig. 3A). We also annotated the genes for their association with neurodevelopmental conditions based on the gene’s inclusion in our curated neurodevelopmental genes set using information from known ASD gene lists^24,27^, ClinGen^47^, and OMIM^48^ (supplementary section 1) and/or in the Genomics England neurodevelopmental disorder recessive genes set^49^, generating a set of 1,309 genes predicted to be associated with neurodevelopmental risk under a recessive model (Fig. 3A; Fig. S1; Table S1).

#### 2.1.2. Minor allele frequency (MAF) thresholds

Because DelCH involves two different variants, the individual frequency of these variants (denoted MAF_Del_ and MAF_SNV_) may be higher than what is typical for pathogenic variants with dominant effects. However, the frequency of their co-occurrence must be small in order for it to potentially have a large effect size. Thus, we calculated the event frequency as the product of the deletion MAF and the SNV MAF (Freq_DelCH_ = MAF_Del_ * MAF_SNV_). We examined DelCH events at different upper bound frequency thresholds, ranging from 0.01 to 0.00005. This approach allows for a lenient inclusion of SNVs and deletions, as long as the product of their MAFs meets the set Freq_DelCH_ threshold for each individual DelCH event. Given the expected stronger deleteriousness of deletions compared to SNVs, MAF_Del_ was capped in keeping with the prevalence of the studied condition. For our study, the maximum value for MAF_Del_ was set at 0.1; a higher MAF threshold would predict homozygous deletions to occur at a frequency above 1%, which would be incompatible with ASD epidemiology^23^.

#### 2.1.3. Identification of DelCH events

A DelCH event requires one allele to be deleted and the other allele to be affected by a functional SNV. Consequently, the deletion and SNV can either be inherited separately from each parent, or one or both can exist only in the child due to *de novo* event(s). For example, the deletion-matched burden analysis (Fig. 2) relies on deletions inherited from one parent, in which case the SNV was either inherited from the other parent or is *de novo*. DelCH events in parents necessarily involve either a deletion transmitted to their children, and therefore a non-transmitted SNV on the remaining allele, or *vice-versa*. Using this framework, we identified a total of 147,737 DelCH events in our dataset (Fig. 3). Due to phasing limitations, only certain subsets of these DelCH events were included in each strategy (supplementary section 2).

#### 2.1.4. Within-family comparison of DelCH events and SNV stratification

The majority of parental couples share the same ancestry (>80% in a 2015 US study^50^ and 94% in the current study). Given that all analytical approaches proposed in this study are based on within-family comparisons, we expect minimal confounding bias conferred by population structure – which includes differences in structural variant haplotypes^51^,- and therefore no correction for population structure is incorporated in our analyses.

For all three approaches, statistical analyses were performed while stratifying by the functional effect of the SNV involved in the DelCH event. Specifically, SNVs were divided into five groups: i) loss-of-function variants (LoF; i.e., stop gain, frameshift indels, and canonical splicing variants), ii) missense variants (i.e., nonsynonymous SNVs), iii) damaging missense variants (i.e., missense variants predicted to be deleterious by at least four out of nine algorithms/conservation scores (SIFT^52^ <0.05, PolyPhen2^53^ ≥0.9, PROVEAN < −2.5, Mutation Assessor^54^ ≥ 1.9, Mutation Taster ≥ 0.5, CADD_phred^55^ ≥15, PhylopMam ≥ 2.3, PhylopVert100 ≥ 4, and within Pfam protein domain with phastcons_placental > 0)^56^, iv) LoF and damaging missense variants combined, and v) synonymous variants. For brevity, we abbreviate the five functional variant types as LoF, Miss, DMiss, LoF_DMiss, and Syn, respectively.

### 2.2. DelCH event comparison strategies

Following these filtering steps, the DelCH events can be compared between cases (children, which may be affected or unaffected) and controls (unaffected parents). We used three different strategies to examine possible association of DelCH with ASD (Fig. 2): (1) “Traditional burden analysis”, (2) “Deletion-matched burden analysis”, and (3) “Transmission Disequilibrium Test” (TDT). For all three strategies, we performed analyses separately for each functional variant type described above, with Syn considered to be a negative control.

#### 2.2.1. Traditional burden analysis: Within-family comparison of DelCH burden between children and their parents

Traditional burden analysis compares the rate of DelCH events between children and their parents (Fig. 2, strategy 1). This strategy includes all deletions (*de novo*, inherited, and non-transmitted). Its main advantage is the inclusion of a larger number of individuals. However, this comes at the expense of a loss of distinction between *de novo* and inherited variants, as well as of phasing information for *de novo* deletions (in children) and non-transmitted deletions (in parents) (see supplementary section phasing considerations). Consequently, when the SNV is outside the deletion boundaries, it cannot be assumed that the two variants are on different alleles (and thus constitute a DelCH event). False positives, i.e., the occurrence of the deletion and SNV on the same allele, could negatively affect the signal to noise ratio in the data. While we recognise such limitation of traditional burden analysis, we estimated the rate of potential false DelCH events in our data at just 4.5% of the total number of DelCH events.

We used logistic regression, stratified by family, to test the hypothesis of an increased burden of DelCH in cases (autistic individuals) compared to controls (parents). Given that each individual presents with a unique set of deletions for this analysis, the totality of sequence on the remaining alleles of deleted regions varies per individual, which affects the *a priori* likelihood of the occurrence of DelCH events - the more sequence is deleted, the higher the probability of additional variation on the remaining allele. Consequently, the *a priori* DelCH probability under the null hypothesis cannot be assumed to be identical between the comparison groups. Thus, covariates for statistical correction in traditional burden analysis included the total length of coding sequence (bp) of all genes affected by deletions, and sex. If data are available for unaffected siblings, the same comparison can be made between parents and their unaffected children.

#### 2.2.2. Deletion-matched burden analysis: A burden analysis of DelCH events in inherited deletions comparing children to their deletion-transmitting parents

The second strategy focuses exclusively on deletions in children that are inherited. The haploid sequence of the remaining alleles of each inherited deletion in all children is queried for SNVs to identify the collective burden of DelCH events. Similarly, the collective burden of SNVs in the haploid sequence of the identical set of remaining alleles in the deletion-transmitting parents is determined (Fig. 2, strategy 2), allowing for a direct comparison between children and deletion-transmitting parents using conditional logistic regression stratified by the deletion. If the data include unaffected siblings, as is the case here, the same analysis can be performed starting with inherited deletions identified in unaffected children.

Note that this approach provides unambiguous allelic phasing; given that the allele with the deletion is transmitted from the deletion-transmitting parent to the child, the remaining allele cannot be shared in this parent-child pair, allowing for the comparison of sequence-level variants in exons of any gene affected by the deletion, both within and outside deletion boundaries (Fig. 1, scenarios 1 and 2, see supplementary section phasing considerations). An additional advantage of this approach is that the location and amount of queried deleted sequence is identical between children and parents, thus the a priori DelCH probability under the null hypothesis can be assumed to be identical between the comparison groups (in contrast with traditional burden analysis).

#### 2.2.3. Transmission disequilibrium test (TDT) of SNVs from non-deletion transmitting parents making up DelCH events in the children

Although traditional burden analysis allows for a larger sample size compared to deletion-matched burden analysis, the potential gain of statistical power is affected by the lack of complete phasing information, which may reduce the signal to noise ratio. To overcome this, the third strategy employs a transmission disequilibrium test (TDT) to examine whether SNVs contributing to a DelCH event are transmitted to autistic individuals at a rate higher than expected (Fig. 2, strategy 3). The vast majority of deletions (97.6%) are inherited, in which case the SNV is transmitted by the parent who does not transmit the deletion. Using Fisher’s Exact Test, the observed transmission rate (OBS) was estimated for a specified type of SNV (“target SNVs”: LoF, DMiss, etc.), while the expected transmission rate (EXP) was estimated on the remaining variant types (“non-target SNVs”). For TDT, homozygous variants in the parents are excluded, as they violate transmission equilibrium. Similarly, scenarios wherein variants are identified in both parents (implying a DelCH in one parent) are also excluded, as this suggests non-pathogenicity of the DelCH event. To boost the statistical power of the TDT, we also incorporate unaffected sibling data, specifically by testing for over-transmission in autistic individuals and under-transmission in unaffected siblings (see equations below).

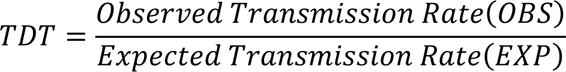

*with:*

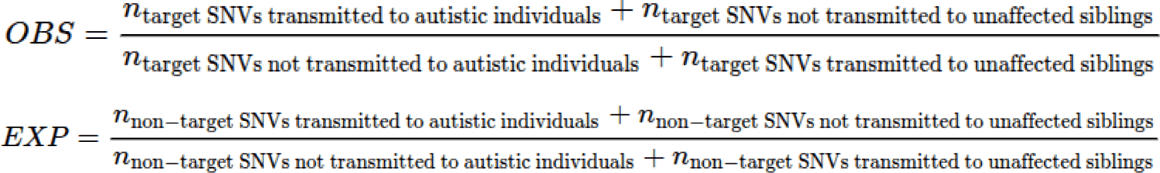

### 2.3. Assessing DelCH in ASD across strategies

To evaluate a potential enrichment of DelCH events in ASD, we calculated the number of genes affected by DelCH in at least two unrelated autistic individuals, while not observed in any unaffected family member. We also calculated the reverse: genes involved in DelCH events in at least two unaffected siblings and not in any individual with ASD. We used permutation to assess statistical significance of any difference between the two results (supplementary section 3).

### 2.4 Gene set enrichment analysis on the candidate genes

To explore common biological pathways among the candidate genes identified from the section 2.3, we used GeneMania to expand the candidate gene list, by 50 genes using information such as gene co-expressions, genetic interactions, and physical interactions, etc.^57^ The different interaction databases were weighted equally in the GeneMania analysis with the option of “equal by data type” selected. Using the extended gene list, we performed gene set enrichment analysis on Gene Ontology gene sets that have the size between 50-2,000 genes using g:Profiler^58^.

## 3. Results

We developed three analytical strategies to investigate the role of DelCH events in disease etiology and implemented these methods using genomic data from ASD cohorts. After data filtering, we identified 147,737 DelCH events in 8,617 families from MSSNG, SSC, and SPARK. Of these, 30,203 events (20%) consisted of SNVs occurring within the deletion boundaries (Fig. 1, scenario 1; Fig. 3B), allowing unambiguous phasing of the SNV and the deletion. In the remaining 117,534 DelCH events (80%), sequencing data from the family members was used to inform phasing of DelCH events where possible (Fig. 3B). Of these, 93,910 were DelCH events with Freq_DelCH_<1% (Fig. 3B). Fourteen percent of deletions involved in DelCH events, are full gene deletion, 46% partially delete at least 10% of the gene body, while 40% overlap less than 10% of the gene body (Table S2).

Given that this dataset included both affected and unaffected children, we performed two iterations of traditional and deletion-matched burden analyses. In the first iteration, we compared the DelCH burden in autistic individuals to that in their parents. As a negative control, we then repeated this approach comparing the DelCH burden in unaffected siblings to their parents. Similarly, we carried out two iterations of TDT, allowing us to compare deviations from the equilibrium in transmissions from parents to their children with ASD versus transmissions from parents to their unaffected children. An overview of our findings is presented in Fig. 4.

**Fig. 4.**
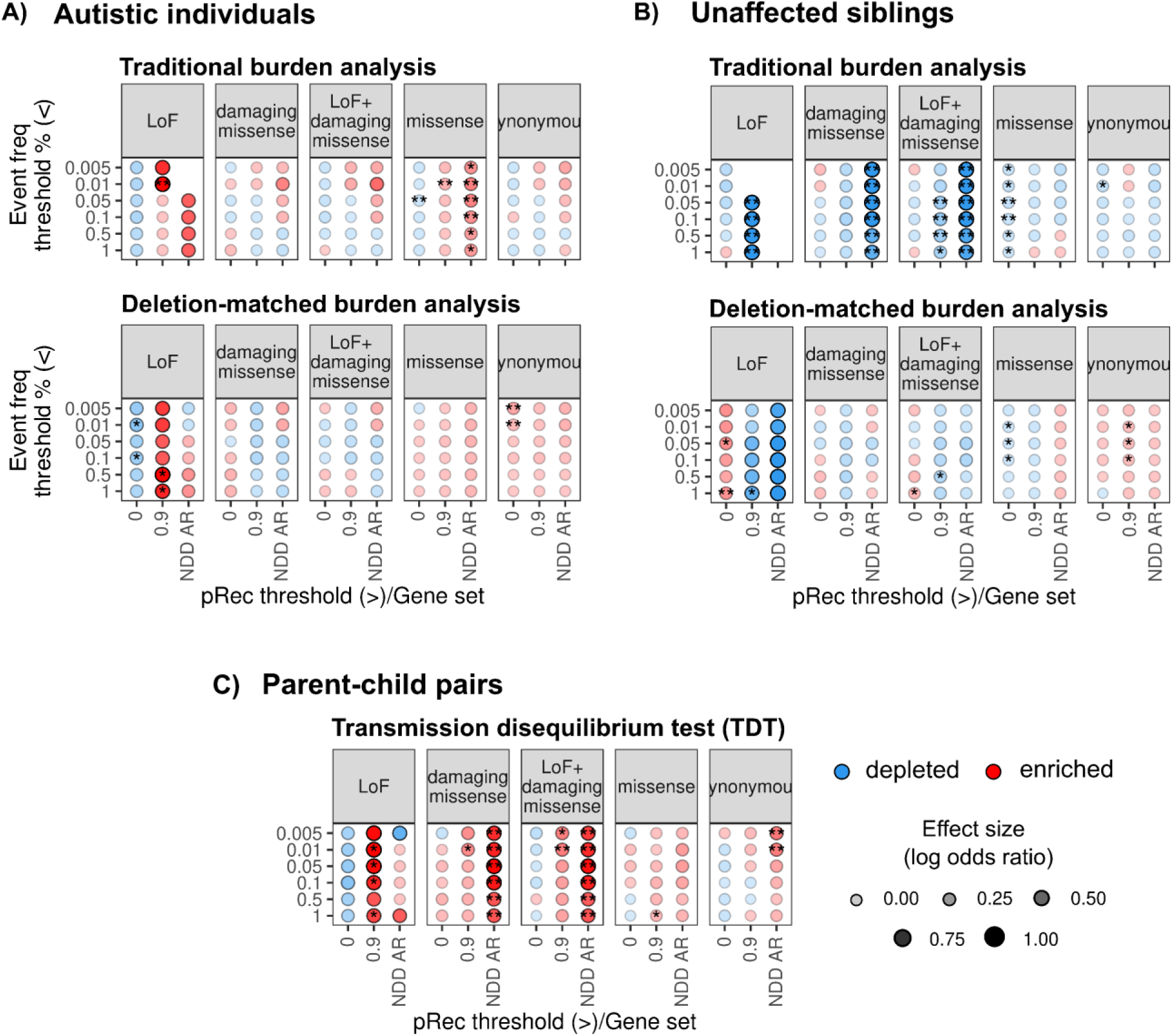
Results from the three proposed strategies in **A)** autistic individuals vs parents, **B)** unaffected siblings vs parents, and **C)** transmission disequilibrium test where we combined the data from autistic individuals and unaffected siblings. Colors indicate the direction of effect of DelCH events, with red indicating enrichment and blue indicating depletion. A single asterisk indicates a trend toward significant with p<0.1, and a double asterisk indicates a nominal significance level (p<0.05). The size of the circles represents the effect size on a logarithmic scale as indicated in the bottom right.

### 3.1. Traditional burden analysis

Comparing DelCH events involving all deletions (i.e., inherited, *de novo*, and non-transmitted) between autistic individuals and parents (significance expressed as one-sided Wald’s p-values, with p < 0.05 depicting nominal significance) we observed an increased burden in autistic individuals for LoF variants in pRec>0.9 genes with marginal p-values at Freq_DelCH_ < 1% (OR=1.59, p=0.050) and <0.5% (OR=1.76, p=0.060).

Unexpectedly, we found a slightly higher burden of DelCH events involving Syn variants in both autistic individual-parent and unaffected sibling-parent comparisons, with ORs range between 1-1.1 in both cases. In our reiteration of the traditional burden analysis using unaffected siblings, we also found a depletion of DelCH events involved LoF and LoF_DMiss in NDD genes and genes with pRec>0.9, although not statistically significant, as well as a nominally significant depletion of DelCH events involved Miss variants at pRec≤0.9 and event frequency cut-offs of 0.1%, 0.05%, and 0.01% (OR ranges between 0.95-0.96).

### 3.2. Deletion-matched burden analysis

Again, all p-values reported in this section are from one-sided Wald’s tests, depicting nominal significance levels uncorrected for multiple testing. We observed a nominally significant enrichment of DelCH events in autistic individuals compared to deletion-transmitting parents in DelCH events involving a LoF or missense variant in genes predicted to be intolerant to biallelic LoF (pRec>0.9) with event frequency less than 0.01% (Odds Ratio;OR_LoF_=2.14, p_LoF_=0.048; OR_Miss_=1.10, p_Miss_=0.043). Limiting the analysis to DelCH events affecting genes in the NDD gene list showed enrichment in autistic individuals of extremely rare DelCH events (frequency < 0.01%) across variant types, but this effect did not reach statistical significance except for missense variants (OR_Miss_=1.33, p_Miss_=0.031). Although not statistically significant, we also observed a general depletion of DelCH events in genes more tolerant to biallelic variation among autistic individuals, across most variant types, except for damaging missense variants, which showed a near-null effect. (OR_DMiss_=1.01) (Fig. 4A). Repeating this analysis in unaffected siblings and their parents, we observed a significant depletion of DelCH events involving DMiss and LoF_DMiss SNVs in NDD genes in any event frequency, as well as LoF and LoF_D_Miss_ in genes intolerant to biallelic LOF (pRec>0.9) with less stringent event frequency cut-offs (Fig. 4B).

### 3.3. TDT

The p-values reported in this section is based on one-sided Fisher’s exact test. We observed a significant over-transmission of variants in DMiss and LoF_DMiss categories involved in DelCH events affecting NDD genes, across all event frequency cut-offs (OR_DMiss_=[1.4-1.76], p_DMiss_<0.05; OR_LoF_DMiss_=[1.37-1.7], p_LoF_DMiss_<0.05). In genes predicted to be intolerant to biallelic LoF (i.e., pRec>0.9), results showed a trend toward significance of over-transmission of LoF (OR=1.77, p=0.076), DMiss (OR=1.2, p=0.089), and a significant over-transmission of LoF_DMiss (OR=1.25, p=0.034) involved in DelCH events with frequency < 0.01%. As a post-hoc analysis, we repeated the TDT analysis but performed it separately for autistic individuals and unaffected siblings. We found that all significant results were not only the result of over-transmission in autistic individuals, but also of under-transmission in unaffected siblings, suggesting negative selection on the involved DelCH events (Fig. S2).

### 3.4. ASD candidate gene prioritization through DelCH events

After identifying genes affected by DelCH events in multiple unrelated individuals, we found seven genes (Table S3) affected by DelCH in two unrelated autistic individuals and no unaffected family members. In contrast, no genes were identified when applying the same criteria to unaffected siblings (compared to all other family members). Results from our permutation analyses indicated a very low probability of finding this case-control difference by chance (one-sided Wilcoxon rank-sum test, p<2.2 x 10^-16^; supplementary section 3; Fig. S3). We examined the DelCH event in the seven genes in the Integrative Genomics Viewer (IGV)^59^ to confirm the CNV and SNV calls for the entire family if the Compressed Reference-oriented Alignment Map (CRAM) file is available (Fig. S5). With IGV visualization, we found the inherited deletion in SP0140142 sample having inaccurate breakpoint. The deletion was no longer overlapping *OR10A7* gene after breakpoint correction. Thus, the *OR10A7* gene was removed from the candidate genes leaving only six genes shown in the Table 1.

**Table 1.** List of DelCH events impacting the six genes affected by DelCH in two unrelated autistic individuals and no unaffected family members. Each unique deletion represents a deletion in one sample and the DelCH event can involve more than one SNV. Remarks; Inh = Inheritance, Freq = frequency, POS = position, M = maternal, P = paternal, D = de novo, A = ambiguous, FD=full-gene deletion, PD=partial-gene deletion (>50% of the gene body is deleted), FS=less than 50% of gene body is deleted but causes a frame-shift, SL=less than 50% of the gene body is deleted but causes a stop-loss.

| Gene | Deletion |  |  |  |  |  | SNV |  |  |  |
| --- | --- | --- | --- | --- | --- | --- | --- | --- | --- | --- |
|  | Coordinate | Size | Inh | Freq | #Genes |  | POS | Inh | Freq | Effect |
| **CFHR4**<br>(chr1) | 196814001- 196936200 (FD) | 122kb | P | 0.04 | 2 |  | 19691290, A-T | M | 0.01 | DMiss |
|  | 196849601- 196915000 (PD) | 65kb | P | 0.03 | 1 |  | 196915074, C-T | M | 0 | DMiss |
| **HSDL1**<br>(chr16) | 83857601-84167800 (FD) | 310kb | P | 2e-4 | 7 |  | 84124642, G-C | M | 0.30 | DMiss |
|  | 84140919-84149519 (PD) | 8.6kb | M | 0 | 2 |  | 84124642, G-C | P | 0.30 | DMiss |
| **MYO15A**<br>(chr17) | 16842001- 18388200 (FD) | 1.5mb | D | 0 | 36 |  | 18153852, A-T | P | 0.42 | DMiss |
|  | 18118994- 18119154 (FS) | 161b | D | 0 | 1 |  | 18153852, A-T | A | 0.42 | DMiss |
| **NEFH**<br>(chr22) | 29485216- 29508910 (PD) | 24kb | P | 0 | 2 |  | 29490053, A-C | M | 0.18 | DMiss |
|  | 29490480- 29497482 (SL) | 7kb | M | 0 | 1 |  | 29490053, A-C | P | 0.18 | DMiss |
| **OR1A2**<br>(chr17) | 3045231- 3621687 (FD) | 576kb | M | 2e-4 | 19 |  | 3198283, G-T | P | 0.55 | DMiss |
|  | 3193869- 3210726 (FD) | 17kb | P | 2e-4 | 1 |  | 3198283, G-T | M | 0.55 | DMiss |
| **OR5P2**<br>(chr11) | 7724378- 7819466 (FD) | 95kb | P | 2e-4 | 1 |  | 7796304, C-G | M | 0.33 | DMiss |
|  |  |  |  |  |  |  | 7796508, T-C | M | 0.33 | DMiss |
|  | 7662601- 7813000 (FD) | 150kb | P | 0 | 3 |  | 7796304, C-G | M | 0.33 | DMiss |
|  |  |  |  |  |  |  | 7796508, T-C | M | 0.33 | DMiss |

Of the six prioritized genes, *OR1A2 and OR5P2* are olfactory receptor genes. Clinically, olfactory disturbances are frequently observed as part of broader sensory abnormalities in many autistic individuals and likely contribute to the commonly observed rigid food preferences^60^. Despite solid evidence for olfactory dysfunction in ASD^61^, the nature of this association remains largely unknown. Possibly, changes in olfactory function may influence social behaviors, and/or variation in some olfactory genes may alter aspects of brain development directly associated with the ASD phenotype. Of note, two other olfactory receptor genes have been associated with ASD, *OR52M1*^62–64^ and *OR2M4*^65^, both of which are strong ASD gene candidates according to SFARI Gene. Of the three olfactory receptor genes prioritized here, *OR1A2* was previously associated with neuroticism^66^.

*MYO15A* encodes myosin, a motor protein involved in actin organization and preservation of cochlear hair cells and their stereocilia ^67^. Autosomal recessive variants in this gene have been associated with hearing loss^68^ and Usher syndrome, while deletions of 17p11.2 involving *MYO15A* cause Smith-Magenis syndrome. Both Usher and Smith-Magenis syndrome are associated with increased rates of ASD and/or autistic features^69,70^. Interestingly, a possible role of *MYO15A* in ASD was previously suggested due to its involvement in a compound heterozygous event identified in monozygotic twins with ASD^71^.

*HSDL1* encodes an inactive enzyme of the short-chain dehydrogenase/reductase superfamily^72^. The protein predominantly localizes to mitochondria, mediated by mitochondrial localization signals encoded in both the amino and carboxyl terminals. However, its biological function in the mitochondria remains to be elucidated. Although HSDL1 lacks enzymatic activity, it is strongly conserved across vertebrate evolution, implying that the protein might perform an essential, but still uncharacterized, process. There is accumulating evidence for a role of mitochondrial dysfunction in ASD^73^.

*NEFH* encodes the heavy chain of neurofilaments—the key cytoskeletal component that provides structural support to axons^74,75^. Both light and heavy chains are potential biomarkers for axonal damage^76–78^, and several studies have reported elevated plasma neurofilament levels in autistic individuals^79,80^. In our analysis, both deletions are short and mainly encompass the C-terminal region of *NEFH*. One of the deletions also includes the last exon of the adjacent gene *THOC5*. Although observed in unrelated individuals, both DelCH events involved the same missense variant (g.22-29490054 A > C; p.E805A), consistent with its high gnomAD allele frequency (0.154). Such a high allele frequency predicts that homozygosity of the missense variant would be expected to occur at a substantial rate in the population and thus is likely benign. We hypothesize therefore that pathogenicity of *NEFH* compound heterozygosity may exist when missense variation on one allele co-occurs with loss of function of the other allele (in this study, a deletion). There are precedents for this mechanism; e.g., congenital sensorineural hearing loss caused by compound heterozygosity affecting *GJB2* involving a rare truncating and a relatively commonly occurring missense variant^81^. Homozygosity of the missense variant can lead to mild to moderate hearing loss but is also found in individuals with normal hearing^82,83^.^81^. Homozygosity of the missense variant can lead to mild to moderate hearing loss but is also found in individuals with normal hearing^82,83^. Autosomal recessive retinal dystrophy can be caused by deep intronic variants in *trans* with a loss of function variant in the *ABCA4* gene^84^. Similarly, compound heterozygosity underpinning oculocutaneous albinism can involve truncating variants in the gene *TYR* in *trans* with missense variants, which by themselves have ∼4% predicted homozygosity in the population^85^.

Finally, *CFHR4* encodes a plasma protein that regulates the complement system and is primarily expressed in the liver^86,87^. Although the two observed DelCH events in our analysis mainly affect CFHR4, one of the deletions also contains the adjacent gene *CFHR1*. Elevated levels of CFHR4 have been reported in individuals with Down Syndrome^88^ and CFHR4 has been identified as one of several immune-related plasma proteins significantly associated with psychiatric disorders in a recent Mendelian randomization study combining proteomics and GWAS findings in schizophrenia, bipolar disorder and depression^89^.

### 3.5 Gene set enrichment analysis shows prominent involvement of neurogenesis

We performed gene set enrichment analysis on the GeneMania-extended network of the six candidate genes and their 50 neighbouring genes (Fig. S6). We identified 32 significantly enriched gene sets (adjusted p<0.05; Table S4) showing a remarkable convergence to processes related to neurogenesis (e.g., neuron and glial cell development, neuron projection, and sensory organ development; Table S4). Out of the 32 enriched gene sets, 13 gene sets involved all candidate genes (either as a membership of the gene set or the member gene is directly connected to the candidate gene). Interestingly, there are common genes linking candidate genes in the network; *GNGT1* links olfactory receptor genes and *CFHR4*, *CDC42* links *CFHR4* and *HSDL1*, *AHI1* links *HSDL1* and *MYO15A*, and *NEFM* and *STXBP1* link *MYO15A* and *NEFH*. This finding highlights how candidate genes with their unique biological function might work closely with each other at higher level in the development of nervous system.

## 4. Discussion

We report three analytical approaches to examining the role of DelCH in disease, and report results from their implementation in a large ASD family dataset. The strongest and most consistent signal across the three methods was observed with LoF variants affecting genes with high predicted intolerance under a recessive model (pRec>0.9). In deletion-matched burden analysis and TDT, these approaches yielded nominal p-values <0.01 in the lower DelCH frequency range (<0.01%), while in traditional burden analysis, similar p-values were recorded in the higher event frequency range (0.5-1%). As shown in Fig. 4, this signal was reasonably consistent across the three methods. Intersecting DelCH events with the *a priori* selected NDD gene set showed enrichment in autistic individuals and depletion in unaffected siblings. While the number of observed DelCH events involving LoF sequence-level variants was too small to allow meaningful observations on individual genes, results from our permutation indicated a very low probability of finding the observed enrichment in ASD by chance (p<2.2 x 10^-16^).

Collectively, our findings indicate a modest enrichment of rare (<0.01%) DelCH events in autistic individuals. Not unexpectedly, DelCH events are rare, requiring large datasets to fully expose their impact on ASD liability. Based on our observations, we conclude that the strongest signals of DelCH events can be expected for LoF variants affecting genes with high predicted intolerance under a recessive model (pRec>0.9). Our *a posteriori* power analysis predicts 80% power to detect significant signals (α=0.05) for variants meeting these criteria with sample sizes of 25,943 and 41,668 for traditional and deletion-matched burden analyses, respectively, and 14,554 families for TDT (supplementary section 4; Fig. S4). Note that our power analysis is specific to ASD, and therefore its applicability to conditions with different genetic architectures may be limited.

Several observations lend circumstantial evidence to our findings: i) genes with high pRec values generated the strongest signals, ii) the enrichment in TDT was observed in LoF and damaging missense variants, but not for missense or synonymous variants, and iii) the observed enrichment of DelCH in autistic individuals did not emerge when traditional and deletion-matched burden analyses were applied to unaffected individuals. Finally, we identified six genes (*CFHR4*, *HSDL1*, *MYO15A*, *NEFH*, and three olfactory receptor genes, *OR1A2*, *OR4P2*) affected by DelCH events in two unrelated autistic individuals, while not observed in non-affected family members. In contrast, applying the same criteria to unaffected siblings, no genes were identified. While evidence linking each of these candidate genes to ASD specifically is tentative, our gene set enrichment analysis strongly suggests that when considered together, they point towards neurogenesis, consistent with our current understanding of the pathophysiology of ASD^90–93^.

Nevertheless, our study has several limitations. First, the ASD dataset examined here is reasonably large in absolute terms but limited in size due to the rarity of the event under study. As a result, our analysis was insufficiently powered to robustly reject the null hypothesis, i.e. the involvement of specific genes in ASD due to DelCH events. In light of the nominal p-values reported for individual comparisons, the prioritization of the seven genes should be considered hypothesis-generating, requiring replication in other studies. Indeed, based on our power analysis, we estimate that sample sizes in the range of 26,000 – 42,000 individuals for burden analyses, and 15,000 families for TDT would be required to address this question with adequate power. Second, it is important to note that while each of the three proposed strategies maximizes advantages of different dataset properties, they are examining the same dataset and therefore are not fully independent tests. The analytical strength of burden analyses lies in the comparison with controls. The strength of deletion-matched burden analysis is the perfect match of queried deleted sequence between cases and controls, whereas traditional burden analysis allows for a larger number of events to be analyzed. However, due to the phasing limitation of *de novo* and non-transmitting deletions, traditional burden analysis may introduce a degree of noise into the data for the case of SNVs outside of the deletion boundaries. To fully benefit from traditional burden analysis, phased genotype data generated through technologies like long-read sequencing is required. Additionally, implementation of deletion-matched burden analysis and TDT in other datasets requires the availability of trio data.

Our present study focused exclusively on DelCH; future work could extend this approach to other types of compound heterozygous events, while taking into account the specific nature of its genomic components. For example, the impact of a duplication fully encompassing a (set of) gene(s) in combination with an SNV/indel on the other allele cannot assumed to be obligatorily deleterious. In contrast, a duplication involving part of a gene may, combined with an SNV/indel on the other allele, may be hypothesized to have deleterious effects similar to that of a DelCH.

In conclusion, findings from our three strategies suggest a modest role of DelCH in ASD and highlight the potential involvement of six genes in ASD under a recessive mode of action. Despite lacking power (the number of families in our analyses was about half of the required number for adequate power), we consistently observed an enrichment of DelCH events in autistic individuals. While individually, our observations were only nominally significant, finding a significantly different number of genes affected at least twice in autistic individuals versus those without ASD provides further statistical support for the hypothesis that DelCH contributes to the genetic etiology of ASD. The analytical pipeline implementing our three complementary strategies for analyzing this specific type of compound heterozygosity is available for application in other studies.

## Data Availability

All data used have been accessible through the 3 databases: MSSNG, SPARK, and SSC autism cohorts. Specific genetic variant events investigated in this study are available as supplementary data also provided with this manuscript.

## Data availability

The 147,737 DelCH events are provided in Table S2. The complete MSSNG and SFARI datasets can be obtained via data access agreements; please see https://research.mss.ng/ and https://www.sfari.org/resource/sfari-base/ for more details.

## Code availability

All the analytical strategies were implemented in R. The scripts developed and used to generate results shown in this study are available on GitHub (https://github.com/naibank/CHASE).

## Author contributions

Conceptualization: JV, BTrost, WE. Methodology: WE, EM, BTrost, JV. Software: WE, KH, RMMWF, SW, FA, AC. Investigation: WE, KH, RMMWF, NBS, DJM, SW, FA, AC, MMA, XZ, RS, NS, JR, MZ. Visualization: WE, KH, RMMWF, NBS. Data curation: WE, NBS, BThiruv, TN, GP, BTrost. Funding acquisition: SWS, JV. Supervision: WE, EM, SWS, BTrost, JV. Writing—original draft: WE, KH, RMMWF, NBS, BTrost, JV. Writing—review & editing: WE, KH, RMMWF, NBS, MMA, XZ, EM, SWS, BTrost, JV. WE and JV have accessed and verified the underlying data. All authors read and approved the final version of the manuscript.

## Acknowledgements

Support from The University of Toronto McLaughlin Centre, the Hospital for Sick Children (SickKids) Foundation, the Ontario Brain Institute, Genome Canada/Ontario Genomics Institute, the Northbridge Chair in Paediatric Research held at the Hospital for Sick Children and University of Toronto (SWS) and the SickKids Psychiatry Associates Chair in Developmental Psychopathology (JV).

