## Supplementary materials for "Evaluating the impact of compound heterozygosity involving microdeletions and sequence-level variants: findings in autism"

##### 1. Generation of our curated neurodevelopmental disorder (NDD) gene list

In order to annotate the genes that are intolerant to recessive variation and are implicated in neurodevelopmental phenotypes, we employed two NDD-relevant gene lists as references. These catalogs include: a curated gene list from Genomics England, which contains putative recessive NDD genes, and a second research-based gene list, which was compiled in our laboratory. For the latter gene list, we attempted to expand the number of genes that are relevant for our research hypothesis with the goal to identify novel gene-disease associations. To determine the genes that were included in our NDD gene list we defined the following criteria. I) Genes reported in the SFARI Gene database and annotated with score 1, 2, 3 or "Syndromic" (SFARI gene list was exported on November 27, 2024)[25.] II) Genes that have been manually evaluated under the EAGLE curation framework, a novel approach designed to assess the level of evidence that associate a gene with ASD risk[22]. EAGLE curated genes classified as definitive, strong, moderate or limited were included in our catalog (annotation was based on the latest update performed in December 2024). III) ClinGen curated genes with a gene-disease validity classification defined as definitive, strong, moderate and limited. Then, we only retained genes that have been evaluated by expert curation panels relevant to neurodevelopmental phenotypes, such as the Intellectual disability and Autism GCEP, or assessed for conditions that include neurodevelopmental symptoms in their clinical profile (The ClinGen list was retrieved on November 27, 2024)[45]. IV) Genes associated with neurodevelopmental phenotypes according to OMIM[46]. To explore the OMIM database, we used a list of 22 search terms associated with different neurodevelopmental phenotypes to compile all possible relevant NDD-genes (OMIM gene lists were retrieved on December 16, 2024). Then, we only retained genes for which the molecular basis of the disorder is known (Phenotype mapping key = 3). Finally, from this NDD-relevant gene list, we filtered the genes based on the probability of being intolerant to biallelic loss-of-function variation (pRec score > 0.9). As a result, we obtained our curated NDD gene list of likely recessive genes relevant to NDD etiology.

<u>OMIM search terms relevant to NDDs</u>
"intellectual disability", "developmental delay", "mental retardation", "cognitive delay", "impaired intellectual development", "global developmental delay", "intellectual impairment", "mentally retarded", "cognitive impairment", "delayed psychomotor development", "psychomotor retardation", "neurodevelopmental disorder", "autism", "epilepsy", "cerebral palsy", "attention-deficit/hyperactivity disorder", "motor disorders", "schizophrenia", "bipolar disorder", "depressive disorders", "anxiety disorders", "obsessive-compulsive disorder"

#### 2. Phasing considerations

Let *D* be a heterozygous deletion affecting one or more coding exons of one or more genes, and let *S* be a heterozygous sequence-level variant (SNV or indel) impacting an exon of one of those genes. Further, let $A_D$ represent the allele on which *D* is found, and similarly for $A_S$. Specifically, if $A_D = M$, then *D* was maternally inherited, and if $A_D=F$, then *D* was paternally inherited. *D* and *S* represent a DelCH event if $A_D \neq A_S$. Below, we describe the analyses presented in this paper and how DelCH events can be identified under various scenarios involving the individuals being compared and the availability of WGS data for those individuals.

##### Strategy 1: Traditional burden analysis - within-family sample-based comparison of DelCH burden

In this analysis strategy, the goal is to compare DelCH burden between children and their parents. The comparisons encompass all DelCH events, regardless of inheritance status. Here, the information available for phasing in the children is not the same as in their parents. Specifically, for the children, we could potentially use parental information to determine whether *D* and *S* are on opposite alleles; however, this is not possible for the parents, since we do not have sequencing data for *their* parents. For this reason, it would create a bias if we used parental information to perform phasing in the children, as we are unable to do the same in the parents. Thus, we do not attempt to perform phasing at all in this analysis—instead, we assume that the presence of a deletion and an SNV/indel constitute a DelCH event.

##### Strategy 2: Deletion –match burden analysis - comparison of DelCH burden between children and deletion-transmitting parents

This analysis strategy involves comparing the burden of DelCH events between children (autistic individuals or unaffected siblings) and their deletion-transmitting parents. For a given child to be included in this analysis, WGS data must be available for that child and at least one of his or her parents. For a given deletion *D* and sequence-level variant *S* as defined above, we can determine whether *D* and *S* constitute a DelCH event as follows.

**Scenario #1: WGS data from both parents are available**. In this case, *D* and *S* can be unambiguously phased using the parental genotypes, regardless of whether *S* is within or outside *D*.

**Scenario #2: WGS data are not available from the non-deletion-transmitting parent**. Suppose that $A_D=M$. If *S* is detected in the autistic individual within *D*, then we know that $A_S=F$. If *S* is outside *D*, phasing may still be possible, depending on the genotype of the mother. Specifically:

- If *S* is present in the mother, then $A_D=A_S$; thus, *D* and *S* do not represent a DelCH event and are not used in the analysis.
- If *S* is not present in the mother, then *S* must have been transmitted from the father and thus $A_D \neq A_S$. (It is also possible, although very unlikely, that *S* could be *de novo*, in which case it could be on either allele). In this case, *D* and *S* represent a DelCH event and would be included in the analysis.

##### Strategy 3: Transmission disequilibrium test of SNVs leading to DelCH events

In this analysis, the starting point is children with exon-affecting deletions. We test whether SNVs affecting genes impacted by these deletions (whether inside the deletion or not) are over-transmitted from the non-deletion-transmitting parent to the child. WGS data from the deletion-transmitting parent are also required in order to ensure that the deletion is not *de novo*.

##### 3. Permutation testing to assess DelCH gene enrichment

We performed permutation testing to assess whether the observed number of genes with DelCH events in ≥2 cases and 0 controls could be explained by chance. Two scenarios were evaluated:

1. Cases: autistic individuals; Controls: all parents + unaffected siblings
1. Cases: unaffected siblings; Controls: all parents + autistic individuals

This design balanced the number of DelCH events in control groups, enabling fair comparison.

We used LoF_DMiss DelCH events from traditional burden analysis as input, where signals were strongest. Of 13,217 such events, 3,765 came from autistic individuals, 1,962 from unaffected siblings, and 7,490 from unaffected family members. Each permutation condition was repeated 100 times with random sampling.

<u>Approach 1: Constant case-to-control ratio</u>

We preserved the observed case-to-control ratio for autistic individuals (3765/9452=0.398) and varied the number of events in cases from 500 to 3,500 in 500-event increments. At each step, we sampled events from both cases and controls, recorded the number of genes with DelCH events in ≥2 cases but absent in controls, and plotted the results (Fig. S3A). The same procedure was applied to unaffected siblings, using both their own (1962/11255=0.174) and the autistic individuals' case-to-control ratio for comparison.

Results showed that autistic individuals consistently had more genes with recurrent DelCH events than unaffected siblings, with the distribution shifting rightward as case events increased. In contrast, distributions for unaffected siblings remained left-skewed. A one-sided Wilcoxon rank-sum test confirmed a significant difference at 2,000 events in cases ($p<2.2x10^{-16}$). Our observed values (seven genes in autistic individuals, none in unaffected siblings) aligned with the distribution peaks, suggesting non-random enrichment.

Approach 2: Constant total number of events

We fixed the total number of DelCH events (7,400 for autistic individuals, 3,800 for unaffected siblings) and varied the proportion of case events from 10% to 50% in 10% increments (Supplementary Fig. 3B). As in Approach 1, we randomly sampled the events according to the specified ratio and assessed how many genes showed DelCH in ≥2 cases but none in controls.

A similar trend emerged: at low proportions, distributions were similar across groups. However, as the percentage of case events increased, the peak for autistic individuals shifted more sharply rightward, indicating greater gene involvement compared to unaffected siblings. This supports the hypothesis that DelCH mechanisms contribute more significantly to ASD than to unaffected status.

#### 4. Power Analysis

A post hoc power analysis was conducted using the pwr package in R to determine the sample sizes required for detecting observed effect sizes at a significance level of $\alpha=0.05$ across varying power levels. Although *a priori* power analysis is preferred, no similar studies evaluating the role of DelCH in ASD exist to our knowledge to provide an estimate of the effect sizes.

Consequently, we performed power analysis using estimated effect sizes from the statistical tests performed in each strategy. For traditional and deletion-matched burden analyses, the null model log-likelihood and the fitted model log-likelihood were extracted from the output of conditional logistic regression (clogit function in the survival package), and used to estimate McFadden's $R^2$. Next, we used the $R^2$ value to calculate the effect size, f2, and fitted an F test that estimates the required sample size using the pwr.f2.test() function. In TDT, we first calculated the proportions of cases and controls carrying DelCH events, then used the proportions to find the effect size, h, using the ES.h() function. Assuming the transmission rate of all SNVs made up of DelCH events is 50%, we conducted a one-tailed two-sample t-test using the pwr.2p.test() function.

### Supplementary Figures

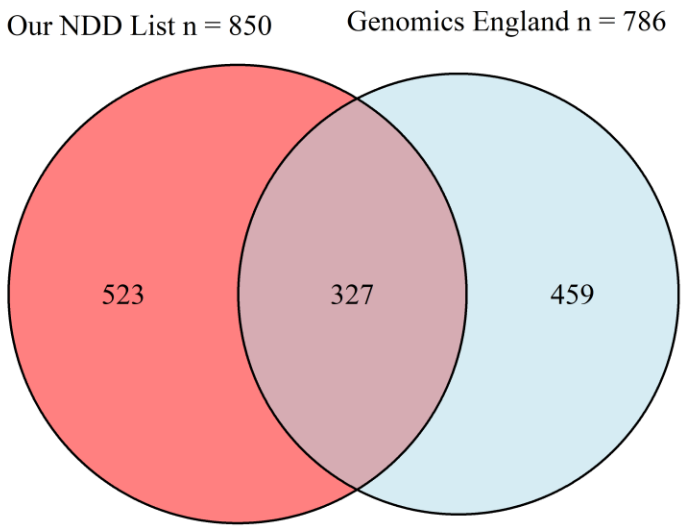


**Fig. S1:** Overlap between Genomics England and our NDD-gene lists. The Venn diagram illustrates the overlap between the common genes included in the curated Genomics England gene list (light blue) and the NDD gene list compiled by our group (red).

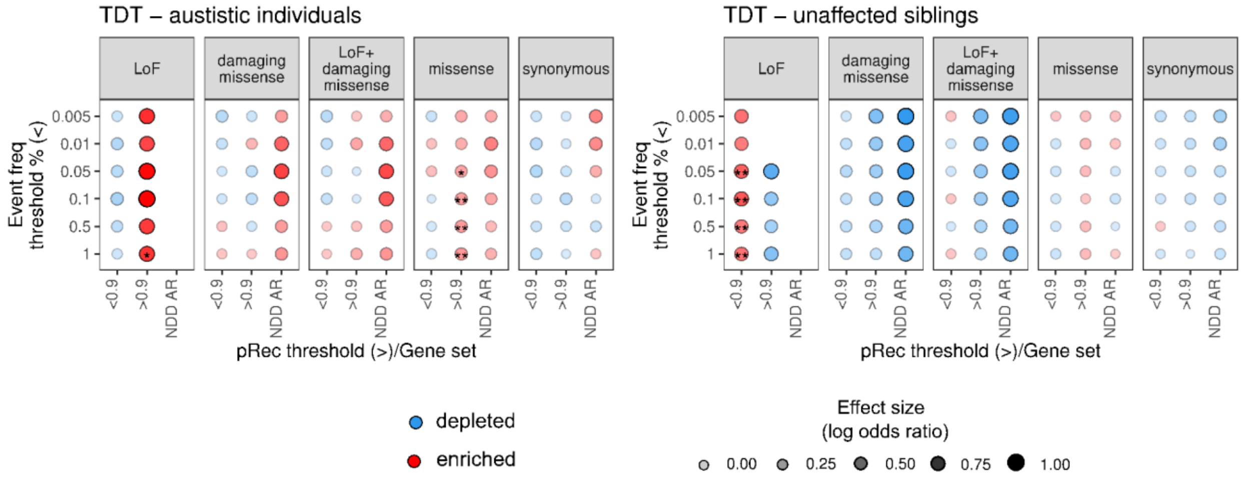

**Fig. S2:** TDT results separating autistic individuals and unaffected siblings. We observed generally an overtransmission of SNVs making up DelCH in autistic individuals, while the opposite was noted in the group of unaffected siblings (i.e., an undertransmission of SNVs). Colors indicate the direction of effect of DelCH events, with red indicating enrichment and blue indicating depletion. A single asterisk indicates a trend toward significant with p<0.1, and a double asterisk indicates a nominal significance level (p<0.05). The size of the circles represents the effect size on a logarithmic scale as indicated in the bottom right.

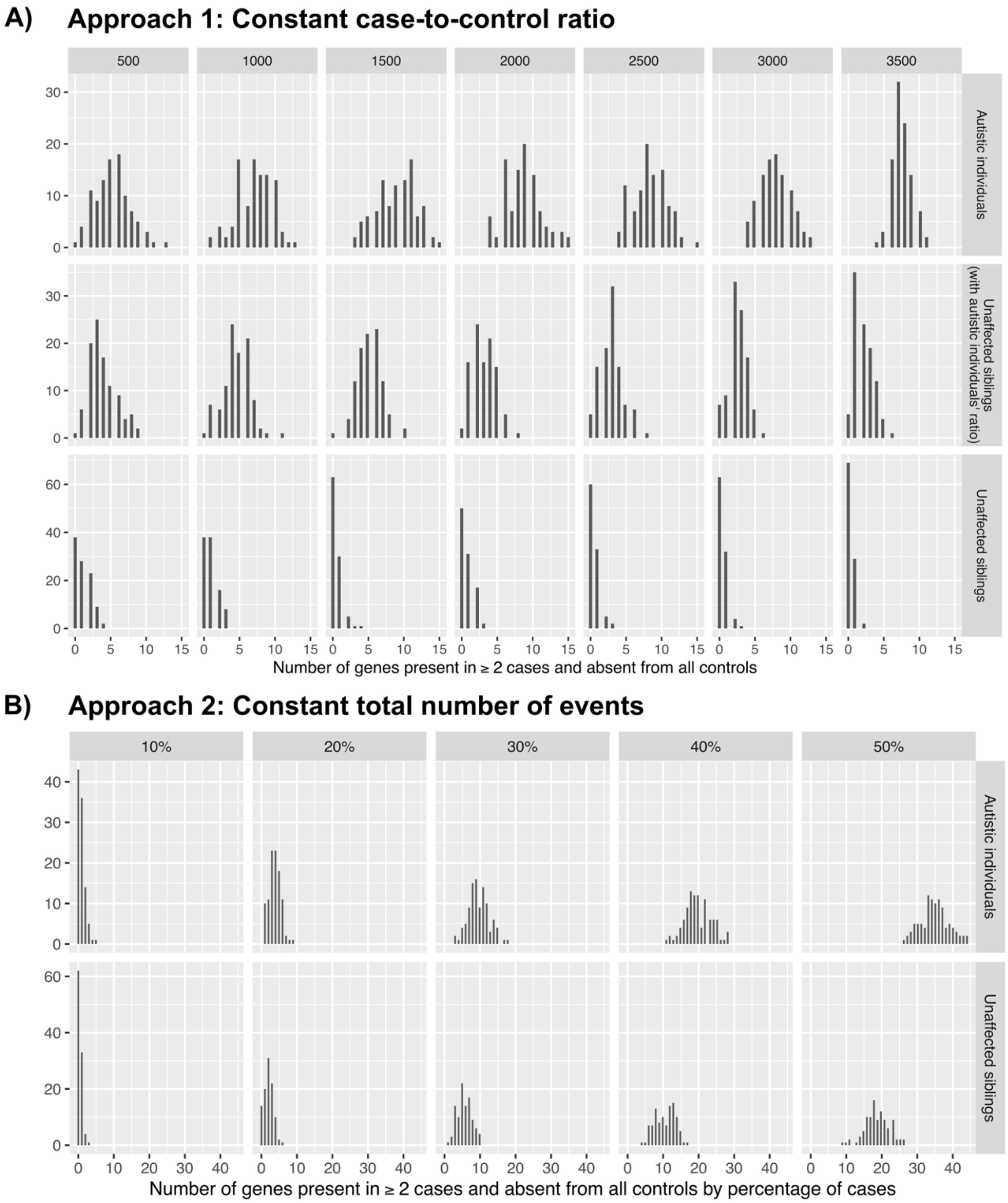

**Fig. S3:** Distributions of genes with DelCH events in ≥2 cases but absent in controls generated by permutations in autistic individuals and unaffected siblings. In **A) Approach 1: Constant case-to-control ratio**, the number of events in cases are varied from 500 to 3,500 in 500-event increments. Note that the events were sampled with replacement for unaffected siblings when n=2000,2500,3000,3500. **B) Approach 2: Constant total number of events** varies the proportion of case events from 10% to 50% in 10% increments. Each permutation condition was repeated 100 times with random sampling.

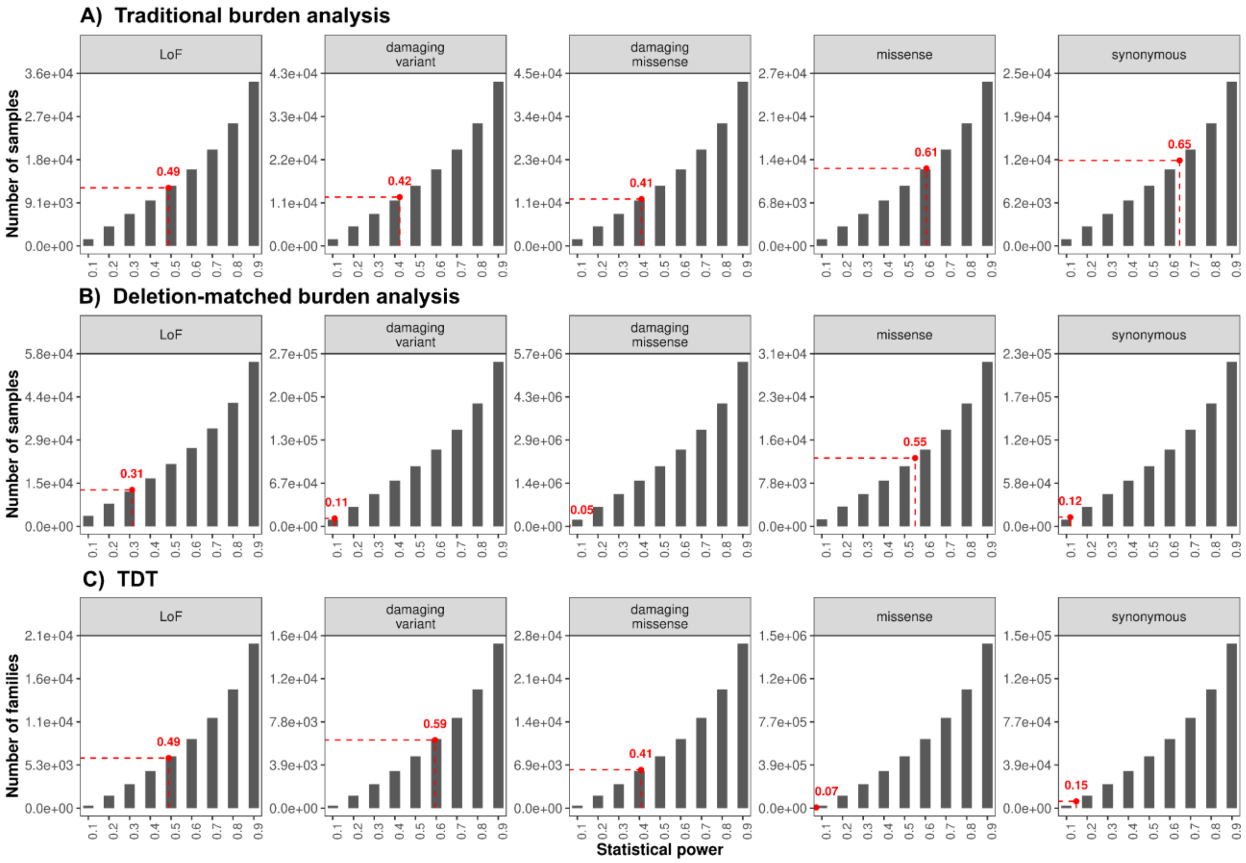


**Fig. S4**: Power analysis for A) **traditional burden analysis**, B) **deletion-matched burden analysis**, and C) **TDT**. The red dotted line indicates the current power.

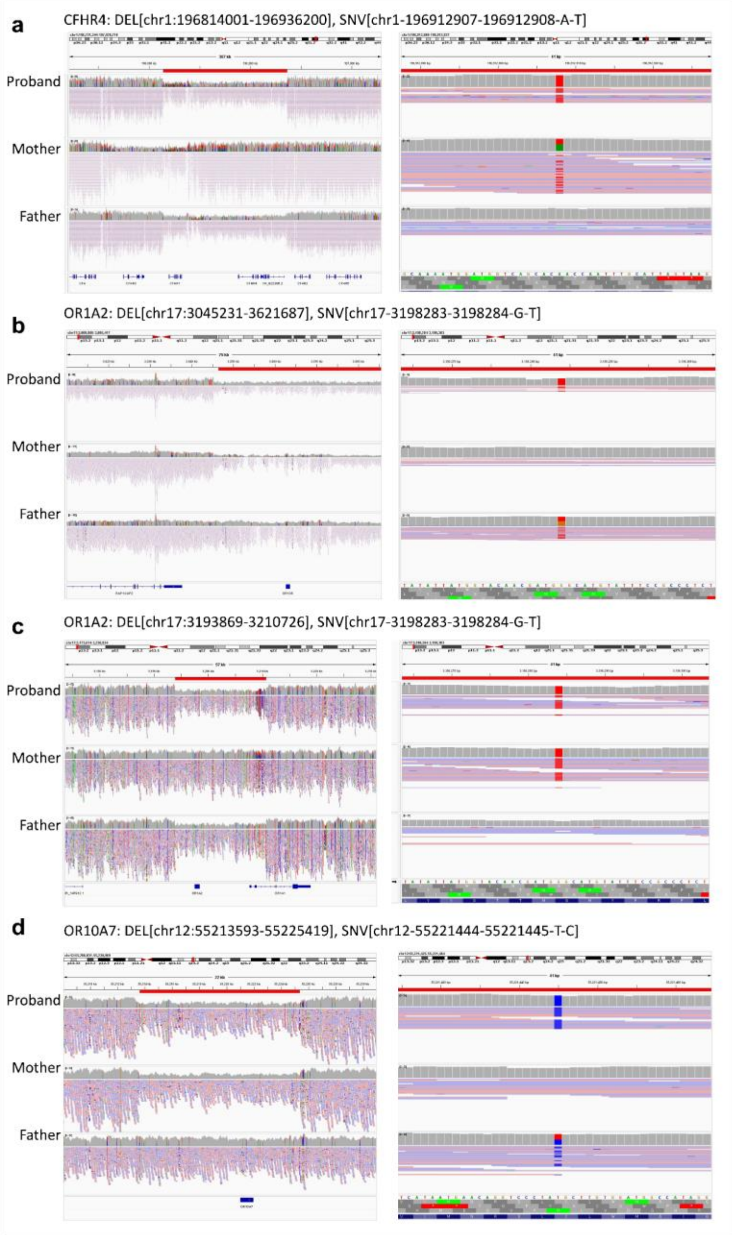


**Fig. S5**: IGV visualization of DelCH events impacting candidate genes. **a)** CFHR4 gene was impacted by a paternal-inherited deletion and a maternal-inherited damaging missense variant in AU3861301 proband. **b)** OR1A2 gene was impacted by a maternal-inherited deletion and a paternal-inherited damaging missense variant in 7-0094-004 proband. **c)** OR1A2 gene was impacted by a paternal-inherited deletion and a maternal-inherited damaging missense variant in 7-0460-003 proband. **d)** OR10A7 gene was impacted by a maternal-inherited deletion and a paternal-inherited damaging missense variant in MSSNG00359-003 proband.

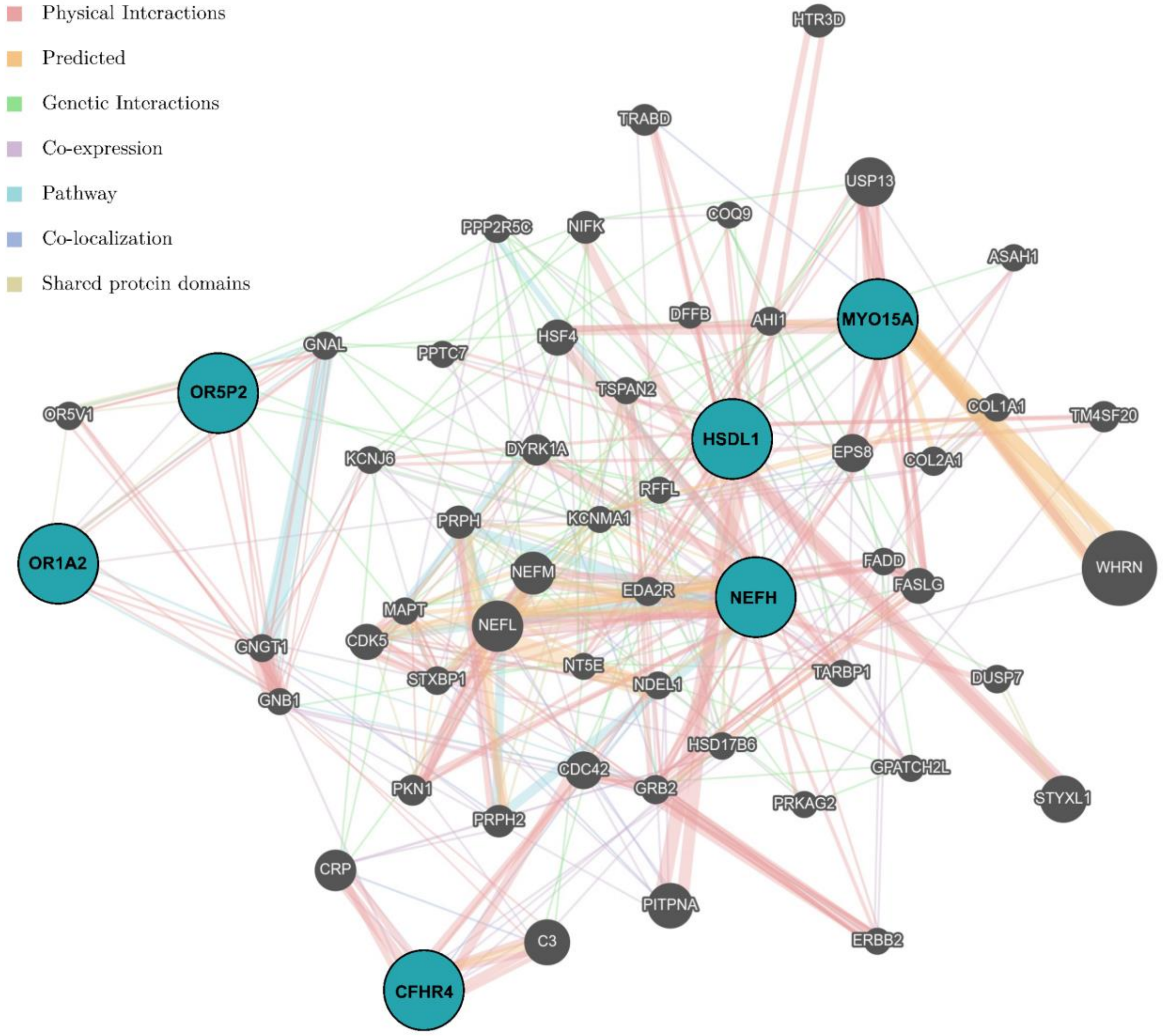


**Fig. S6:** GeneMania-extended gene network of six candidate genes and their 50 closest neighbouring genes. The candidate genes with DelCH events identified in autistic individuals but not in the unaffected family members, are highlighted in teal circles. Size of the node indicates strength of genes being relevance to the network, while the size of the edge represents the strength of the link between genes. Edge color indicates the type of interaction.
